# Correlates of Self-Sampling Willingness for HPV-DNA Testing among Medically Underserved Rural Kenyan Women: A Mixed Methods Study

**DOI:** 10.1101/2024.09.21.24313929

**Authors:** Joyline Chepkorir, Nancy Perrin, Lucy Kivuti-Bitok, Joseph J Gallo, Deborah Gross, Jean Anderson, Nancy R Reyolds, Susan Wyche, Hillary Kibet, Vincent Kipkuri, Anastacia Cherotich, Hae-Ra Han

**Author notes:** **Corresponding author:** Joyline Chepkorir.

## Abstract

**Introduction:** Cervical cancer is the leading cause of cancer-related deaths among Sub-Saharan African women, particularly in rural areas where screening rates are lower due to limited access to highly sensitive tests. This study aimed to investigate factors that might influence rural Kenyan women’s willingness to self-collect samples for HPV-DNA testing.

**Methods:** This study utilized data from a mixed-methods study in Bomet and Kericho Counties, including survey responses from 174 women and semi-structured interviews with a subset of 21 participants. Logistic regression was used to analyze quantitative data and theoretical thematic analysis for qualitative data.

**Results:** The surveyed women had a mean age of 45.2 years, were mainly uninsured (76%) and from low-income households (88.4%). Most participants had heard of cervical cancer (83.2%), yet only 6.4% had ever been screened. However, 76.9% expressed willingness to self-collect samples for HPV-DNA testing. Factors significantly associated with increased self-sampling willingness were cervical cancer awareness (OR=3.49, 95% CI=1.50-8.11), having health workers (OR=1.88, CI=1.23-2.86) and the news media (radio and television) (OR=2.63, CI=1.27-5.48) as primary sources of health information, and ever hearing about cervical cancer from the news media (OR=2.43, CI=1.07-5.51). Conversely, high cervical cancer stigma (OR=0.71, CI=0.57-0.88) and longer travel time of 30 to 120 minutes to the nearest health facility (OR=0.44, CI=0.20-0.93) were associated with decreased willingness. Interview data corroborated these findings.

**Conclusions:** Cervical cancer screening uptake is notably low among rural Kenyan women in Bomet and Kericho Counties. Sample self-collection for HPV-DNA testing appears widely acceptable. A comprehensive approach involving educational outreach, health worker recommendation, and mass media campaigns could enhance cervical cancer screening via self-sampling, potentially reducing the burden of cervical cancer. Future research should employ implementation science methodologies to explore cervical cancer screening uptake via self-sampling, to inform population-based implementation strategies in Kenya.

## Introduction

Cervical cancer (CC) is the most common cancer among women in Sub-Saharan Africa, particularly among women aged 21 to 48 years.^1,2^ In Kenya, CC is the leading cause of cancer-related deaths, with about 5,236 new cases diagnosed annually.^3^ Over 90% of women are diagnosed at advanced stages despite prior contact with the healthcare system.^4^ About 63% of invasive CC cases in the country are caused by high-risk human-papillomavirus (HPV) types 16 and 18, which can be prevented through HPV vaccination.^3^ A two-dose HPV vaccination program was introduced in 2019, targeting adolescent girls and young women in schools and health facilities.^5^ Unfortunately, the HPV vaccine coverage remains low as only 31% of the targeted group had received the two doses of HPV vaccine by 2021.^5^

CC screening is a promising strategy to detect CC among an estimated 13 million Kenyan women aged 15 years and older, who are at risk for the disease.^6^ Kenya’s National Cervical Cancer Screening Guidelines have been implemented for over a decade and currently recommend screening of all eligible women aged 30 to 49 years old. ^7^ As part of the 2030 global cancer elimination strategy, the World Health Organization (WHO) recommends a target screening of 70% of eligible women and treatment of 90% of those diagnosed with CC in each country.^8^ However, the screening uptake rate is approximately 16 to 18% in screening-eligible Kenyan women aged 18 to 69 years and the rates are notably lower in rural areas.^4^ The WHO and Kenyan cancer screening guidelines recommend HPV-DNA testing as the primary screening method since it has a high sensitivity to detect high-risk HPV sub-types.^9,10^

In Kenya, over 90% of health facilities use visual inspection methods and a few of them use Pap smear and HPV testing, which are often constrained by sociocultural beliefs, limited availability of skilled workforce, and essential infrastructure.^11^ Self-sampling approach for HPV-DNA testing offers an opportunity to circumvent these barriers by paving the way for community-based screening, increased reach and empowerment of women to collect specimen in private spaces at their convenient place and time; without fear of pain associated with pelvic examinations used in other procedures.^11^ Nonetheless, the success of self-sampling depends on the target women’s acceptability.^12^ Although studies in Sub-Saharan Africa have affirmed a high acceptability of self-sampling in diverse settings, few studies have investigated the acceptability of this procedure in rural community settings.

Empirical studies on the use of self-sampling in rural communities in Kenya are rare. In a qualitative study conducted among rural Kenyan women (N=120), women reported generally positive experiences with HPV self-sampling. In an urban sample (N=409), more than 80% of women reported that they would be comfortable using a self-sampling device and 84% would prefer at-home sample collection.^12^ Similarly, another study among rural Kenyan women (N=97) found that 90% of them were willing to collect their samples in private places.^6^ As HPV testing and self-collection kits gradually become available in Kenya’s health care system, it is critical to understand factors that may impact women’s willingness to self-collect samples for HPV-DNA testing.^9^

### Purpose

This study’s purpose was to identify potential barriers and facilitators to self-sampling, to inform the design and implementation of interventions aimed at promoting women’s adoption of self- sample for HPV-DNA testing. To the best of our knowledge, factors associated with HPV-DNA self-sampling willingness have not been investigated in rural, community-based contexts in Kenya and other parts of SSA.

## Methods

### Theoretical Framework

This study was guided by the socio-ecological framework, which proposes that an individual’s behavior affects and is affected by the social environment.^13^ It emphasizes the need for health interventions to target changes at the individual/intrapersonal, interpersonal, organizational/institutional, community, and policy levels to support and maintain healthy behavior.^13^ We presupposed that at the individual level, an individual’s willingness to screen might be influenced by their capacity to make health care decisions, and access to resources (i.e. health insurance), their knowledge and awareness of cervical cancer (i.e. need for screening), health literacy.^14^ At the interpersonal level, we theorized that an individual’s self-sampling willingness could be influenced by interactions with their peers and household dynamics (i.e. spouse’s involvement in a woman’s healthcare decision-making).^15^ At the organization level, we presumed that health workers and the health facility level could potentially impact a woman’s willingness to self-collect samples for HPV testing.^16^ At the community level, we hypothesized that factors such as beliefs and CC stigma might influence self-sampling willingness.^17^ Lastly, certain health system and policy-level factors such as the geographical location of health facilities have the potential influence successful self-sampling.^15,16^

### Study design and setting

This study was a secondary analysis of data from a parent convergent mixed-method study, which explored the interplay of health information, health literacy, and cervical cancer screening uptake. The study used interviewer-administered surveys and semi-structured interviews, to collect data between August and September 2023 in rural Bomet and Kericho Counties, Kenya. Kenya is subdivided into 47 Counties. Bomet and Kericho Counties, located in Kenya’s South Rift of the Great Rift Valley, consist of pre-dominantly rural populations, 96.8% and 89.6%, respectively.^18^ These Counties are adjacent and consist primarily of Kipsigis communities that have comparable cultural and economic practices.

### Study Sample

Women were recruited from community settings. Convenience sampling was used to select voluntary participants for the quantitative arm, who were available and met the inclusion criteria. Eligibility criteria were 1) residing in rural Kericho or Bomet County, 2) female aged 18 to 65 years (age range for CC screening based on Kenya’s guidelines), 3) proficient in Kipsigis language, 4), had educational attainment of grade 8 or lower. Participants were excluded if they were either 1) acutely/terminally ill, or 2) were cognitively impaired, limiting participation in study activities. A total of 174 eligible women participated in the survey study. Purposive maximum variation sampling was used to select a sub-sample of participants (n=21) with varied characteristics for semi-structured interviews.^19^ Specifically, CC screening status (ever versus never screened) 45 years), and anticipated CC stigma (none versus at least one) were used as the basis for selection for qualitative interviews.

### Procedures

The survey, interview guide, and consent form were originally developed in English, translated to Kipsigis, and then back translated by experts to ascertain the accuracy of the translation.^20^ Two bilingual registered nurses from each County and one medical doctor specializing in obstetrics and gynecology reviewed the survey and interview guide for face validity and contextual relevance. Based on their feedback, we removed some questions that were not contextually relevant and added questions assessing risk factors for CC. The survey was pre-tested among the first 24 women while the interview guide was pilot tested among 10 women. Amendments to the survey and interview guide were minor and mainly involved the re-phrasing of questions for contextual aptness.

Five trained research staff residing in the two Counties recruited study participants, using study flyers and word of mouth, from participants’ homes, local health centers, shopping centers, and churches. The interviewer-administered survey and interviews were offered in convenient private spaces of the participants’ choosing (e.g. inside or outside their house). The survey was administered on RedCap software offline mode due to limited access to the Internet and immediately transferred to the web after data collection. All interviews were audio-recorded to facilitate verbatim transcription and analysis. To maintain participants’ confidentiality, all audio recordings were exported to a secure platform before deleting from the original devices. Each survey and interview took approximately 30 to 45 minutes. Participants were incentivized with Ksh.330 (approximately $2.37) for responding to the survey and additional Ksh.330 for those who were selected for interviews. Upon completion of data collection, transcripts and survey data were checked for accuracy and completeness and cleaned as needed.

### Ethics

Study procedures were approved by **(blinded for review)** Institutional Review Board **(# blinded for review).** Additionally, in line with the local requirements for conducting human subjects research, study procedures were reviewed **(blinded for review)** and licensed (l**icense No. Blinded for review)** by the Kenya National Commission for Science, Technology and Innovation. The research staff obtained verbal consent in Kipsigis language from each participant before the study activities. The consent document detailed information about the study’s purpose, risks, and benefits of participating. Each participant could ask questions about the study procedures before consenting.

### Measures Independent variables

Factors at the individual level assessed were demographics including age (in years), marital status (single, married, separated, divorced), education (no formal education, between grade 1 and 3, between grade 4 and 8), employment (self-employed, unemployed, employed in private sector), health literacy level, perceived health status (poor, fair, good, very good), and CC awareness (ever/never heard about CC). Additionally, resources considered were household monthly income $(less or equal to 35, 36-70, 71-106, 107-142, 143-178, 179-214, 215-251, 252-322), income comfortability (comfortable, or not comfortable), and health insurance status (insured or uninsured), primary sources of health information (news media, social network and community leaders, health workers, teachers and herbalists), and estimated travel time to the nearest health facility (in minutes). Each of these variables were included in the study questionnaire as individual items except for health literacy which used a validated instrument.

The Health Literacy Test for Limited Literacy (HELT-LL) was used to assess participants’ health literacy skills.^21^ The tool, consisting of 12 items, was developed, and tested in a sample of South African women (with high school levels or lower education), who were recruited from primary health clinics in two semi-rural towns.^21^ The internal consistency reliability of the HELT-LL, measured using Cronbach’s alpha, was 0.6.^21^ In our study sample, the internal consistency reliability assessed using Cronbach’s alpha was 0.49. To improve the reliability of the scale, we removed 4 items with item-total correlations less than 0.49. Items removed were questions assessing three dimensions 1) numeracy skills (prescription medication schedule and blood pressure reading), 2) communicative/interactive skills (one’s frequency of asking questions from a nurse/doctor/pharmacist), and 3) electronic health literacy skills (one’s capacity to use of cellphone or computer to answer health-related questions). The items left on the final instrument used for this sample assessed numeracy-, print-, critical-, and oral/communicative health literacy skills. The reliability of these items was 0.57, which is within the satisfactory range. ^22^ Items were scored on a scale of 0 to 1; with possible total scores ranging from 0 to 8.

Participants in this study with scores less than the mean (4.2) were deemed to have inadequate health literacy and those with scores greater or equal to the mean were regarded as having inadequate health literacy levels.

Factors considered at the interpersonal level in this study were: healthcare-decision making (self, self and spouse, spouse only, or mother]), interpersonal sources of CC information (social networks, community leaders, health workers, teachers), and interpersonal sources of general health information (health workers and social networks). Participants could select more than one response. At the organizational level, factors examined were the reliance on health care workers as primary sources of health information (doctors, nurses and community health workers), receiving CC-related information health care workers (whether they had ever heard about cervical cancer from health workers) and prior CC screening (ever screened, or never screened). Factors assessed at the community level were the reliance on the news media (TV and radio) as primary sources of health information, having heard about CC information from the news media and anticipated CC stigma. The 8-item CC stigma scale was used to determine anticipated CC stigma.^17^ Lastly, at the health system and health policy levels the ranking of the nearest health facility (level 1 to 4) and travel time to the nearest health facility (in minutes) were evaluated.

### Dependent variable: Self-sampling willingness

Willingness to self-collect samples for HPV-DNA testing was assessed using a two-part question. The first part of the question explained what the self-sampling procedure is, “HPV self-sampling allows women to collect their own samples for cervical cancer screening using a swab or a brush”. The second part assessed willingness to accept self-collection. “Would you be willing to self-collect a sample for testing if this screening is suitable for you?”. Responses were binary (yes/no).

### Data analysis

The analytical sample consisted of participants (n=24) who were involved in the pre-testing of the survey and 150 women who completed the study survey. Analysis of quantitative data used descriptive statistics (means, standard deviations, medians, range, frequencies, and percent) to summarize the sample characteristics. Also, we used binary logistic regression to assess the correlates of self-sampling willingness. We excluded one survey participant with a missing response on self-sampling willingness; hence the logistic regression output is based on 173 respondents. Quantitative data analysis was conducted in STATA/BE version 17 software.

Qualitative data were analyzed using Dedoose software, in Kipsigis language, by four bilingual coders. We used theoretical thematic analysis to identify codes and generate themes relating to study objective. ^23^ Additionally, we employed inductive analysis by allowing the research objective to evolve through the coding process.^23^ Specifically, each transcript was coded by two coders independently and all coders met regularly to resolve any discrepancies and discuss emerging codes. Coded files were exported in Word format. Themes, sub-themes, and accompanying quotes were organized in an Excel spreadsheet before being translated to English by two of the coders. Thereafter, results from each study arm were merged and juxtaposed using a joint display.^24^

## Results

Table 1 summarizes the characteristics of the survey sample. The mean age of the survey sample was 45.2 (SD=13.2) years. The majority were from Bomet County (64.4%). More than half (81.5%) of the sample had attained formal education between grades four and eight and the majority were married (83.8%). About 78% of the women were self-employed. Most participants (88.4%) earned $35 or less per month in their households. The sample was predominantly uninsured (76%), and the majority (63%) rated their health status as either good or very good.

**Table 1:** Sample characteristics (n=174)

| Characteristic | n (%) / mean (sd) / median (range) |
| --- | --- |
| Age (Mean (SD)) | 45.3 (13.2) |
| Marital status |  |
| Married | 145 (83.3%) |
| Unmarried | 29 (16.7%) |
| County of residence |  |
| Bomet | 112 (64.4%) |
| Kericho | 62 (35.6%) |
| Education |  |
| No formal education | 32 (18.4%) |
| Grade 1-3 | 52 (29.9%) |
| Grade 4-8 | 90 (51.7%) |
| Employment status |  |
| Self employed | 134 (77%) |
| Unemployed | 34 (19.5%) |
| Employed in private/public sector | 5 (2.9%) |
| Missing | 1 (0.6%) |
| Household monthly income |  |
| ≤ (35 USD) | 154 (88.5%) |
| 36 -142 USD | 20 (11.5%) |
| Insurance status |  |
| Insured | 41 (23.6%) |
| Uninsured | 131 (75.3%) |
| Missing | 2 (1.1%) |
| Health status |  |
| Poor/Fair | 65 (37.4%) |
| Good /Very good | 109 (62.6%) |
| Health literacy |  |
| Adequate | 64 (36.8%) |
| Inadequate | 110 (63.2%) |
| Nearest Health Facility |  |
| Level 2 (dispensary/clinic) | 132 (75.9%) |
| Level 3 (health center) | 23 (13.2%) |
| Level 4 and 5 (county hospital/referral) | 19 (10.9%) |
| Transportation to nearest health facility |  |
| Walk | 136 (78.2%) |
| Motorcycle | 35 (20.1%) |
| Public vehicle | 2 (1.2%) |
| Missing | 1 (0.6%) |
| Travel time to nearest health facility |  |
| <30 mins (reference) | 76 (45%) |
| 30-120 mins | 93 (55%) |
| Missing | 4 (2.3%) |
| Health care decision-making |  |
| Self | 139 (79.9%) |
| Self and Spouse | 12 (6.9%) |
| Mother or spouse | 7 (4%) |
| Missing | 16 (9.2) |
| Primary sources of health information* |  |
| News media (tv and radio) | 120 (69%) |
| Social networks and community | 64 (36.8%) |
| Health workers | 121 (69.5%) |
| Other (herbalist, teachers) | 2 (1.1%) |
| Sources of CC information* |  |
| News media (tv & radio) | 64 (37%) |
| Social networks | 36 (20.8%) |
| Healthcare workers | 42 (24.3%) |
| Other (teachers, religious leaders) | 3 (1.7%) |
| CC awareness |  |
| Ever heard of CC | 144 (82.8%) |
| Never heard of CC | 29 (16.7%) |
| Missing | 1 (0.6%) |
| CC screening status |  |
| Never screened | 163 (93.7) |
| Ever screened | 11 (6.3%) |
| Anticipated CC stigma |  |
| Yes | 101 (58.1%) |
| No | 73 (42%) |
| Willingness to self-sample for HR-HPV DNA testing |  |
| Yes | 133 (76.4%) |
| No | 40 (23%) |
| Missing | 1 (0.6%) |

Key: sd – standard deviation \* Select all that apply responses

Most participants had inadequate health literacy levels (63%). The nearest health facilities accessed and used by most participants (75.7%) were level 2 (dispensary/clinic) and most of them walked (78.5%). Travel time to the nearest health facilities was between half an hour to two hours for many (45%) of them. Approximately 80% of participants made their own healthcare decisions.

Regarding health information access, about 69.4% and 70% of participants reported that they primarily accessed health information from health workers and the news media, respectively. Most participants (83.2%) had heard about CC from the news media (37%), their social networks (20.8%), health workers (24.3%), and/or other sources (1.7%). Only 6.4% of the sample had ever been screened for the disease.

The CC stigma scale showed that half of the participants endorsed between one and eight potential drivers of stigma. Only 1.7% of participants perceived they were at risk for cervical cancer. Most participants, 76.9%, reported that they would be willing to self-collect samples for HPV-DNA testing if offered. Among women (n=40) who were unwilling to self-collect samples, the majority (72.5%) indicated that they lacked the confidence to collect samples, while others reported that they were not interested (4%), and afraid of either the procedure (3.5%) or the results (0.6%). Sample characteristics for the sub-sample of participants interviewed are summarized in Appendix A.

### Factors associated with self-sampling willingness

Bivariate logistic regression outputs of sample characteristics on self-sampling willingness are summarized in Table 2.

**Table 2:** Bivariate analysis of sociodemographic characteristics on self-sampling willingness (n=173)

| Characteristic | OR | P-Value | CI |
| --- | --- | --- | --- |
| Age (Mean (SD)) | 0.98 | 0.14 | 0.95-1.01 |
| Marital status | 0.68 | 0.47 | 0.24-1.93 |
| Education | 1.06 | 0.78 | 0.68-1.68 |
| Employment Status | 1.04 | 0.91 | 0.49-2.24 |
| Income | 0.94 | 0.67 | 0.71-1.24 |
| Comfortability with income | 0.57 | 0.33 | 0.18-1.78 |
| Insurance status | 1.35 | 0.50 | 0.56-3.21 |
| Health status | 1.71 | 0.06 | 0.98-3.00 |
| Healthcare decisions | 1.07 | 0.84 | 0.51-2.27 |
| Cervical cancer awareness | 3.49 | 0.004** | 1.50-8.11 |
| Health literacy | 0.99 | 0.99 | 0.80-1.24 |
| Prior cervical cancer screening | 1.38 | 0.69 | 0.29-6.66 |
| Cervical cancer stigma | 0.71 | 0.001** | 0.57-0.88 |
| Nearest health facility | 0.96 | 0.86 | 0.58-0.59 |
| Distance to nearest health facility |  |  |  |
| <30 mins (ref) |  |  |  |
| 30-120 mins | 0.44 | 0.032* | 0.20-0.93 |
| Transportation to nearest health facility | 0.64 | 0.24 | 0.31-1.34 |
| Sources of cervical cancer Information |  |  |  |
| News media (tv & radio) | 2.43 | 0.03* | 1.07-5.51 |
| Social networks | 0.61 | 0.24 | 0.27-1.38 |
| Healthcare workers | 1.48 | 0.41 | 0.59-3.7 |
| Primary sources of health information |  |  |  |
| News media (tv & radio) | 2.63 | 0.01** | 1.27-5.48 |
| Social networks and community | 1.06 | 0.77 | 0.72-1.55 |
| Health workers | 1.88 | 0.003** | 1.23-2.86 |
| OR - odds ratio, CI – confidence interval, SD – standard deviation, ref - reference<br>*- p-value<0.05, ** - p value ≤0.01 |  |  |  |

Results indicated that those who had heard about CC were 3.49 times more willing to self-collect samples than those who had not (odds ratio [OR]=3.49, 95% confidence interval [CI]=1.50- 8.11). Respondents whose sources of cervical cancer information were the news media (radio and television) were 143% more likely to accept self-sampling if offered (OR=2.43, 95% CI=1.07-5.51). Using the news media and health workers as primary sources of health information was associated with 163% (OR=2.63, 95% CI=1.27-5.48) and 88% (OR=1.88, 95% CI=1.23-2.86) higher odds of self-sampling willingness.

On the other hand, cervical cancer stigma was associated with 30% lower odds (OR=0.71, 95% CI=0.57-0.88) of self-sampling willingness. Traveling for half an hour to two hours, compared to less than half an hour, to the nearest health facility was associated with 56% lower odds (OR=0.44, CI=0.20-0.93) of self-sampling willingness. Appendix B summarizes the socio-ecological barriers to self-sampling willingness based on the results.

### Qualitative findings

Qualitative findings were mainly congruent with quantitative results. Nonetheless, we identified some discrepancies in qualitative results; some participants who had heard about CC from the news media were unwilling to self-collect samples for HPV testing. Additionally, while most participants anticipated CC from their community, some participants reported that stigmatizing attitudes from people may be buffered by Christian religious beliefs. Below are the key themes and exemplar quotes:

#### Cervical cancer awareness

Women who were agreeable to self-sampling demonstrated that their willingness was potentially influenced by their understanding of the significance of screening for CC. Some participants who expressed interest in self-sampling desired more education on the procedure prior to collecting the sample. One respondent in her 60s mentioned that she had heard that it is important to detect the disease at its early stages as it allows for early treatment.

*“I will agree (to self-collect a sample for screening). It was said that it is good to screen so that it (cervical cancer) can be prevented before it gets worse.”*

#### Acquisition of health information from health workers

Respondents whose primary sources of health information were health workers (doctors, nurses and community health workers) highlighted the significant role of health workers in promoting knowledge on screening. Also, among some participants trust in health care providers, in addition to health care provider recommendation were facilitators to self-sampling. One participant in her 40s affirmed her willingness to self-collect a sample for HPV-DNA testing:

*“I will not refuse especially when a healthcare provider advises, because I also want to know my health status*.”

*The role of the news media sources*.

Qualitative data showed that the news media promote CC knowledge and could influence self- sampling willingness. One participant in her 30s who had heard about CC mentioned that she became aware of the need for women getting screened through an educational program on the radio, where she primarily accessed health information.

*“I heard a talk on cervical cancer on the radio. Otherwise, I have not seen (the disease); I only hear.”*

However, there were some women who were unwilling to self-collect samples for screening despite demonstrating good knowledge and awareness of screening through the news media. A participant in her 40s mentioned that she had learned the need for women to screen for CC, but she was unwilling to self-collect a sample.

*“I have heard (about cervical cancer) from an educational program on the radio. They made an announcement that women should go to the hospital for screening. The radio is where I primarily get (cervical cancer information) they always teach, emphasizing that one should go for* ***screening.”***

#### Anticipated cervical cancer stigma

Several women from both counties anticipated CC stigma in their community, mainly attributed to the symptoms of the diseases. However, some women indicated that religion might reduce instances of CC stigma. For instance, a participant in her 50s divulged that CC symptoms might drive a woman’s spouse to infidelity, leading to distress and self-blame on the woman.

*“ Cervical cancer might cause a woman to produce discharge with a foul odor. Her husband*

*might reject her and go find another woman. This would cause distress to the woman because she sees that the disease led to infidelity Some men would reject their wives when they are diagnosed with this disease because of lack of sexual intimacy.”*

Contrary to some observations of stigma, some women revealed that individuals with religious beliefs might not stigmatize a woman diagnosed with cervical cancer. A participant in her 50s stated that a woman with cervical cancer might be stigmatized by some of her friends who are non-believers.

*“Friends who are religious will give her hope. Those who have secular beliefs might say she got the disease because of infidelity.”*

#### Longer travel time to the nearest health facility

Several participants reported that they had not been screened due to their inability to cover transportation costs to the health facilities and lack of screening equipment in accessible health facilities. Others reported that accessible health facilities did not have the necessary screening equipment.

*“Nothing prevented me from (going for screening) except for transportation costs. (There was) a free mass screening campaign, but it was announced a day before the screening date. ”*

Some participants disclosed that their nearest health facilities were not well equipped to conduct cervical cancer screening.

*“It (the health facility) is poorly equipped, there is no screening equipment. There are no machines to be used in screening.”*

### Integration

Table 3 below is a joint display illustrating the integration of mixed methods results. For each measure, we provide recommendations for successful adoption of self-sampling for HPV-DNA testing, based on the merged findings. These recommendations accentuate the need to 1) address the significance of CC screening during CC awareness campaigns, 2) optimize patient-provider interactions to promote HPV testing via self-sampling, 3) promote health worker-led CC information dissemination, 4) leverage mass media to boost CC education and screening efforts, 5) complement radio broadcasts of CC education and screening, with health worker-led interventions, 6) encourage discussions about CC in communities to debunk stigma, 7) explore the potential mediating role of religious leaders in reducing CC stigma, 8) increase awareness of CC symptoms and the availability of treatment, 9) create support groups for CC patients, and 10) provide HPV testing kits in community settings to promote access and increase screening uptake among eligible women.

**Table 3:** A joint display of qualitative, quantitative and mixed methods meta-inferences of self-sampling willingness.

|  | Quantitative Findings | Qualitative Findings | Mixed Methods Meta-Inferences |
| --- | --- | --- | --- |
| Measure | OR (95% C.I.) | (Themes and Quotes) | Recommendations for enhancing self-sampling for HPV-DNA testing |
| Cervical cancer awareness | 3.49 (1.50-8.11) | Knowledge about the significance of screening: *"I had heard the health care workers saying that it is necessary for women to get screened so it can be known if we have disease in our bodies, to get regular screening."* (between 20 and 25 years, willing to self-collect a sample). | Address the significance of screening in CC awareness campaigns, to promote screening through sample self-collection for HPV-DNA testing. |
| Acquisition of health information primarily from health workers | 1.88 (1.23-2.86) | Trust in health care providers: *"I trust a doctor or a nurse. you know, they trained in the health care field and they explain things about health and wellbeing very well, based on what you tell them. When you discuss your health issue with them, they would give you a detailed explanation about it."* (between 46 and 50 years, willing to self-collect a sample, primarily accesses health information from a doctor or a nurse) | Optimize provider-patient interactions to educate patients on the importance of CC screening and offer sample self-collection kits to women who are eligible and willing to screen. |
|  |  | • Cervical cancer information acquisition from health workers: *"I have come to know that it (screening) is important because cervical cancer has caused the death of so many women. When health workers came (to our community) to educate people (about cervical cancer screening), I understood that it is important for one to go for screening, so that if you have it (cervical cancer), you can be treated when curative treatment is still possible."* (heard of CC, between 51 and 55 years old, willing to self-collect sample, primarily accesses health information from a doctor or a nurse). | • Promote health worker-led CC information dissemination to ensure provision of accurate CC information and sample self-collection for HPV-DNA testing, among eligible women. |
| News media sources of cervical cancer information | 2.63 (1.27-5.48) | • Television and radio sources: *"I have heard (cervical cancer information) over the radio.... and watched on the television."* (between 61 and 65 years, willing to self-collect a sample) | • Maximize the potential for these mass media to facilitate CC information dissemination and screening uptake. |
|  |  | • Radio-based educational programs and announcements: *"I have heard from the radio...It was said that it affects women, particularly the cervix...It was announced that screening was being done for free at the hospital."* (between 56 and 60 years, unwilling to self-collect a sample, primarily obtains health information from the radio, heard of CC) | • Complement radio-based CC information dissemination in local dialects with education and support from health workers. |
| Anticipated cervical cancer stigma | 0.71 (0.55-0.88) | Community: *"you know if someone has cervical cancer, when they have reached the late stages, there is nothing to hide. It can be concealed when they are still alive but when they die, they you will hear that they had this type of cancer."* (between 36 and 40 years, willing to self-collect a sample) | Encourage discussions about CC as a disease to normalize the topic and enhance screening uptake. |
|  |  | stigma and potential mediating role of religion: *"If it is a Christian family her spouse might not stigmatize her. He will take care of her. But if it is a place where there is no salvation, it is possible that he might stigmatize her and even send her away because there is nothing else going on."* (between 30 and 35 years, willing to self-collect a sample) | Explore the potential role of religious leaders to lead open dialogue about CC and reduce stigma associated with CC. |
|  |  | Community stigma: (a woman with cervical may be stigmatized in the community) *"...because cervical cancer is a bad disease, and it was said that it causes one to have a foul odor."* (between 45 and 50 years, willing to self-collect a sample) | Increase awareness of CC symptoms and treatment, and create support groups for those affected by CC. |
| Longer travel time to the nearest health facility | 0.44 (0.20-0.93) | Travel distance barriers: (I have never gone for screening because the health facility). *"...where screening is being conducted is far."* (between 51 and 55 years, willing to self-collect a sample) | Provide self-sampling kits in community-based settings to ensure easy access ad screening uptake |

## Discussion

We sought to investigate barriers and facilitators of rural Kenyan women’s willingness to adopt self-sampling for HPV-DNA testing. The sample predominantly consisted of women with low CC screening uptake, most of whom expressed willingness to self-collect samples for CC screening if offered. Among the factors investigated, CC awareness, acquisition of health information primarily from health care providers and the news media and having heard about cervical cancer from the news media were positively associated with self-sampling willingness. In contrast, CC stigma, and traveling for 30 minutes or more to the nearest health facility were negatively associated with self-sampling willingness. Of the total sample, 76.9% stated that they would self-collect samples for CC screening if offered. This is consistent with findings from a longitudinal study that sampled rural Malawian women (N=122); 66% of them expressed willingness to self-collect samples for HPV testing.^25^ This study, however, found differences between hypothetical willingness to self-collect samples and uptake of self-sampling. Only 53% and 47% of those who were either willing or unwilling to self-collect samples for HPV testing at baseline, respectively, self-collected samples 12 to 18 months later.^25^ While findings from both studies illustrate generally high proportion of women willing to self-collect samples for HPV testing, they accentuate the need to understand the factors that influence uptake of sample self-collection for HPV testing.

CC awareness and women’s willingness to screen for CC are key in the prevention of HPV and CC.^26^ In this study sample, 83.2% of women had heard of CC, but only 6.4% had ever been screened for the disease in their lifetime. These findings closely align with results from a different rural sample in Uganda, which found that a high proportion of women (82%) had heard about CC awareness yet only 6% had ever been screened for pre-cancerous lesions. CC awareness was significantly associated with 249% increased likelihood of self-sampling willingness. These findings emphasize the need to provide information on CC and self-sampling. Prior studies have established that awareness of screening methods and place of screening are critical in women’s use and access to CC screening services.^20,27^ Qualitative findings demonstrated that primary prevention through education on the importance of CC screening and a demonstration of self-sampling are key facilitators to self-sampling willingness. Comparably, researchers investigating a sample of Ugandan women found that self-sampling acceptability was higher when providers educated women and allowed them to examine the brush before self- collection.^28^ Prior research has shown that compared to urban women, rural women in SSA have considerably lower CC awareness and knowledge, mainly due to constrained access to CC information.^20,29^ To encourage sample self-collection for HPV-DNA testing among women, CC education including practical instructions on self-collection will be essential.

Our findings highlight the potential instrumental role of the traditional news media, mainly the radio, in enhancing HPV-DNA testing. Women whose primary source of health information, and those who had heard about CC from the news media were more willing to self-collect samples. Our results are consistent with prior research findings which reported that women in SSA often learn about CC from televisions, radios.^30–33^ Among rural Tanzanian women, the odds of screening were increased by about 25% among those who regularly listened to the radio.^34^ While the news media play a critical role in CC information dissemination, qualitative findings showed a lack of self-efficacy to self-collect samples for screening might be a hindrance to women who are willing to screen. Stakeholders should ascertain the level of understanding of self-sampling procedure among women who are willing to screen.

Healthcare workers (HCWs) are critical players in the dissemination of information on CC and CC screening. Women who primarily acquired health information from HCWs had 88% likelihood of self-sampling willingness. Qualitative findings indicated that many women learned about CC from HCWs, and some reported that they would most likely accept self-sampling if recommended by a healthcare provider. Although we did not find any significant association between having heard about CC from HCWs and self-sampling willingness, previous research findings showed that receiving information from healthcare providers on CC and CC screening were significantly associated with higher odds of screening uptake.^32,35^ Health care providers, particularly nurses who mainly run rural-based health clinics that were used by the majority of the study sample, should not only offer CC education in health facilities but also conduct community outreach in rural regions that are farther away from these facilities. CC education should be culturally sensitive, elaborate on CC screening procedures, and emphasize the significance of screening, to promote screening uptake.^29^

Prior research findings have established that cervical cancer stigma is a key barrier to disclosure of symptoms, seeking services for CC screening, and treatment of CC, in many Sub-Saharan African communities.^12,36–38^ In this study sample anticipated cervical cancer stigma was high, with 57.8% of the sample endorsing at least one anticipated cervical cancer stigma. This is higher than was found in another Kenyan rural sample of women (n=419) in which only 20.3% anticipated cervical cancer stigma.^36^ The difference might be attributed to sample characteristics as the study recruited women from health facilities; 55.6% of whom were HIV positive, while this study recruited women predominantly from community settings and only 2.5% were HIV positive. HIV positive women possibly have more opportunities to interact with healthcare providers who may provide support and information on CC, ultimately reducing stigma. To promote cervical cancer screening in rural communities, it would be important to destigmatize CC in both community settings and health facilities.

Longer travel time to the nearest health facility was significantly associated with a 56% decrease in the odds of willingness to accept self-sampling in this study sample. Indeed, previous research studies conducted in various countries in SSA indicate that perceived problems with long travel time and high costs associated with long distances to health facilities are barriers to CC knowledge and screening.^38–40^ In a pooled sample of women (n= 40,555) included in the Demographic and Health Surveys conducted between 2013 to 2021 in Benin, Cote d’Ivoire, Cameroon, Kenya and Namibia, 62.4% of women who reported that distance to health facilities was a big problem were rural residents.^40^ About 8% (n=12,899) of all women who indicated that distance to health facilities was a big problem in the study had been tested for cervical cancer compared to 13.5% of women who did not have a problem.^40^ Expanding the quantitative findings, qualitative findings showed that the high cost of transportation to more equipped health facilities was the main reason why women were concerned with travel time and distance, which correspond to results from other studies in Kenya and other parts of SSA.^12,20,38^ Level II health facilities were the most accessible to our study sample. These facilities are financed by the County governments and even though they usually offer free CC screening, conducted by nurses, using VIA/VILI techniques, supplies may be out of stock and participants who are willing to screen may need to seek CC screening services at more advanced health facilities.^36^ To promote self-collection of samples for HPV testing, ministries of health in Kenya and other parts of SSA should ensure that self-sampling kits are readily accessible and available in rural health facilities.

## Limitations

This study has some limitations. First, convenience sampling was used to select participants. This type of sampling is prone to selection bias. Second, data collection was limited to rural-based women from two Counties and findings may not be generalizable to other settings. Third, data were self-reported; it is possible that recall bias may have led to participants under-reporting or over-reporting on some variables. Lastly, we found some discrepant results from qualitative interviews, hence further studies are needed to support our findings. Nonetheless, a key strength of this study is the availability of both qualitative and quantitative data, which facilitated integration and an in-depth understanding of the sociodemographic factors that could impact women’s self-sampling willingness.

## Conclusions

In summary, self-sampling for HPV-DNA testing is a promising approach to screening and early detection of CC among rural Kenyan women. To enhance successful adoption by women, targeted CC education, leveraging the news media and health workers will be instrumental in the improvement of CC knowledge and debunking of stigma among women and consequently their willingness to self-collect samples for HPV-DNA testing. Besides, the implementation of strategies to promote access to screening among medically underserved rural Kenyan women will be of the essence.

## Author contributions

Conceptualization, J.C., N.P., and H.-R.H.; quantitative data analysis, J.C., and N.P. Qualitative data analysis, J.C., A.C., H.K., and V.K. Writing original draft, J.C., and H.-R.H. Abstract and full article review, J.C., N.P., L.K.-B, D.G., J.J.-G., J.A., N.R.-R., S.W., A.C., H.K, V.K., and H.-R.H. All authors have read and agreed to the published version of the manuscript.

## Data Availability

https://datadryad.org/stash/share/fjdv_UKmPJbViLbFkEUxegSJFjIcewyuZBFVmPhiK8s

## Acknowledgements

We would like to extend our gratitude to all the participants who contributed their time and insights to this research study.

## Notes

### Competing Interest Statement

The authors have declared no competing interest.

### Funding Statement

This research received funding from the National League for Nursing, USA, and the Johns Hopkins University School of Nursing (Discovery and Innovation Award).

### Author Declarations

Johns Hopkins Medicine Institutional Review Board

